# Evaluating the acceptability, usability and clinical appropriateness of *Your Path*, an AI-powered tool facilitating relevant access to HIV services post-HIV self-testing in South Africa

**DOI:** 10.1101/2025.11.10.25339576

**Authors:** Nomsa B. Mahlalela, Onthatile Maboa, Caroline Govathson, Laura Rossouw, Lawrence Long, Ross Greener, Sarah Morris, Shawna Cooper, Sasha Frade, Sophie Pascoe, Candice Chetty-Makkan

## Abstract

Timely linkage to HIV prevention and treatment services following HIV self-testing (HIVST) remains a challenge in many countries. While HIVST offers privacy and convenience for individuals to know their HIV status, many do not engage in follow-up care due to behavioural barriers like uncertainty about next steps, stigma, and privacy concerns. This study evaluated the acceptability, usability, and clinical appropriateness of *Your Path*, an Artificial Intelligence (AI)-powered tool designed to facilitate linkage to appropriate HIV services following HIVST. This study was conducted with community members (CMs) and healthcare providers (HCPs), recruited from a research site in an urban area in South Africa. CMs were randomly assigned a mock HIV test result and completed a pre-test survey assessing intentions to seek HIV services, followed by a simulated HIVST session guided by *Your Path*. A post-test survey then evaluated changes in intention to seek care (a proxy for uptake of care) and a System Usability Scale (SUS) measured usability of the tool. A sub-sample of 25 CMs completed in-depth interviews exploring their experience with *Your Path*. HCPs reviewed transcripts of CM tool interactions to assess clinical appropriateness, completeness and relevance of the generated summaries. Qualitative data were analysed thematically, and results aligned with the Theoretical Framework of Acceptability (TFA). Of the 100 enrolled CMs, 59.0% were female and 51.0% received a mock HIV-negative result. After interacting with *Your Path*, 91.0% of CMs reported that it positively influenced their intention to access HIV services. The tool demonstrated high usability, with a mean SUS score of 81.6 (SD 17.5). Most found *Your Path* easy to use (83.7%) and did not need assistance (76.4%), and 94.9% expressed willingness to use it again. However, 15.3% noted that it did not always provide consistent responses to questions and that responses varied or were unclear. CMs described *Your Path* as a helpful guide with a user-friendly design. Most appreciated its private, non-judgmental tone, which helped reduce stigma during interactions. Some valued the sense of confidentiality offered by the tool, while others expressed concerns about the protection of their information. HCPs found the conversational summaries clinically appropriate, and cited the tool as informative and beneficial for supporting linkage to HIV care. Limitations in emotional responsiveness and potential language barriers were noted. Recommendations included integrating local languages, offering data-free access, and aligning with existing healthcare systems to improve reach and coordination of care. *Your Path* was acceptable, usable, and clinically appropriate for supporting linkage to HIV services after HIVST, and showed potential to support intentions to seek HIV care. Larger in-field studies are needed to understand the implementation, cost and sustainability of digital interventions in public health.

## INTRODUCTION

The global HIV/AIDS epidemic continues to be a major public health challenge, with an estimated 44.1 million deaths since the start of the epidemic [1]. Transmission persists globally, affecting all countries including South Africa. Undiagnosed HIV significantly contributes to the ongoing transmission of the virus, and early detection remains key to reaching epidemic control. HIV testing is a critical step in diagnosis and linking infected individuals to effective HIV treatment and care, and those at increased risk of acquiring HIV to prevention services. Early treatment initiation among those infected with HIV has been shown to reduce rates of HIV transmission, improve health outcomes and reduce the cost of care [2, 3, 4]. To increase access and uptake of HIV testing, South Africa offers extensive HIV testing programmes, with HIV self-testing (HIVST) included as a supplementary strategy in the National HIV Testing Services Policy in 2016, especially among key and under-tested populations [5].

HIVST is available in many countries, including South Africa, offering individuals a discreet and convenient method to learn their HIV status. However, many individuals who self-test do not proceed with confirmatory testing if the result is positive as required by the World Health Organisation (WHO) [6,7], or access preventive services such as pre-exposure prophylaxis (PrEP) if the result is negative. Limited linkage to care following HIVST is often due to uncertainty about next steps, stigma and discrimination, fear of negative attitudes from healthcare providers, concerns about privacy, or structural barriers such as cost and time constraints [8, 9]. Effective linkage to confirmatory testing, HIV treatment, care, and prevention, including PrEP, is therefore crucial following an HIVST particularly those conducted in home settings where direct support and referral from healthcare providers is not immediately available.

Artificial intelligence (AI) is evolving to support health decision-making and presents potential to address behavioural barriers to accessing appropriate HIV care and prevention services. In healthcare settings, the transformative role of generative AI models are increasingly being recognised for their ability to enhance health education, deliver perceived empathetic support, and manage data [10]. For instance, large language models (LLMs) including ChatGPT and Claude can engage with users by answering health-related questions, offering tailored guidance, and facilitating linkage to care using de-stigmatising language [11]. These tools can support clients through sensitive health journeys such as HIV testing and treatment initiation, especially where stigma may otherwise hinder engagement [12].

More broadly, AI applications are being leveraged to strengthen public health responses. For example, predictive analytics is being used to forecast disease outbreaks, identify high-risk populations, and inform targeted interventions and resource allocation [13, 14]. In Kenya, AI tools have been deployed across 1800 healthcare facilities to enhance HIV case finding and tailor prevention strategies to individuals at highest risk [15]. AI tools that integrate computer vision and conversational agents are also being scaled up across sub-Saharan Africa to support HIVST by enabling real-time interpretation of test results and facilitating reflexive linkage to appropriate services [16, 17]. These innovations demonstrate how AI can help close gaps in the HIV care continuum by offering more responsive, scalable, and client-centred approaches. However, despite this momentum, the use of AI-powered conversational agents specifically to support linkage to HIV prevention or treatment following HIVST remains limited in low and middle-income countries (LMICs) where the burden of HIV is highest. Given South Africa’s high HIV prevalence, widespread mobile phone access, and evolving digital health infrastructure, there is a critical opportunity to evaluate the potential of AI-enabled tools to improve linkage to care and prevention services following HIVST in ways that are private, personalised, and scalable.

Building on *Your Choice*, an earlier AI-powered tool for HIV risk assessment and personalised prevention guidance, *Your Path* was developed to engage users in post-HIVST conversations via an AI-powered interactive WhatsApp based chat. This study evaluated the acceptability, usability, and clinical appropriateness of *Your Path* in facilitating engagement with HIV services following a simulated HIVST in South Africa.

## METHODS

### Study design

This exploratory evaluation using quantitative and qualitative data collection procedures was conducted with community members (CMs) and healthcare providers (HCPs) recruited from a behavioural research hub in an urban area of South Africa. The behavioural research hub focuses on testing behavioural interventions for HIV prevention and care among adult CMs and HCPs [18]. Qualitative findings are reported following the Consolidated Criteria for Reporting Qualitative Research (COREQ) guidelines (Online Resource 1).

### Recruitment

CMs and HCPs were contacted telephonically to gauge interest in the study and to verify their eligibility. Only CMs (≥18 years) who were willing to provide written informed consent in English, self-reported an HIV-negative or unknown status, and able to travel to the study site were invited and eligible to participate. Those who self-reported as HIV-positive were excluded. HCPs who were willing to provide consent in English and had at least one year of experience in HIV service provision were also invited to participate. The study was conducted with 100 CMs and 25 HCPs in private rooms in healthcare and research settings. During the data collection, only the researcher and the research assistants were present.

### Study procedures

Eligible CMs were randomly assigned a picture of a mock HIV-negative or positive test result, and asked to envision themselves in that scenario when interacting with the tool. This approach was used to simulate real-world post-HIVST decision-making and explore how the tool might influence care-seeking intentions in both outcomes. CMs completed a pre-test baseline survey (Online Resource 2) which included validated Likert scale items to assess intentions to engage in HIV care [20]. CMs who received a negative mock result answered questions about their intention to access HIV prevention and future treatment services, if needed. Those who received a positive mock result answered questions about their intention to access HIV treatment services. CMs were then provided with study phones and instructed to open the *Your Path* thread on WhatsApp by typing “hi” which triggered the welcome message and the menu options. From the main menu, they were instructed to select the “Self-test support” option and complete a simulated HIVST using a WHO-approved oral fluid HIV self-test guided by *Your Path*. Upon completion of the simulated self-test, CMs initiated a chat with the *Your Path* AI-Companion discussing their simulated (mock) test results. The *Your Path* AI-companion aimed to simulate a real-world post HIV test counselling session by providing personalised guidance and support to access HIV services. Chats occurred in a private space to ensure participant comfort and confidentiality. To further protect confidentiality, CMs were asked not to enter any personal information. All interactions were conducted using study access accounts and these interactions were logged and recorded for analysis.

After engaging with *Your Path*, CMs redid the intention to engage in HIV care survey (Online Resource 3) and completed the validated Systems Usability Score (SUS) [19], to measure changes in intention to seek care and usability of *Your Path*. A subset of CMs were purposively selected for semi-structured interviews to explore the acceptability of *Your Path* features, as well as barriers and motivators to engagement. To allow for variation, CMs were purposively selected based on age, gender and mock test result (negative or positive). The interviews were audio recorded and included prompts (Online Resource 4) regarding the CM’s experience using *Your Path* and suggestions for improvement. Each interview lasted about 20 minutes and was conducted in a private room where only the participant and the interviewer were present. The interviews were conducted by two female study team members (NM and OM) with Masters degrees who are experienced researchers. The sample size was adequate to see a recurrence in themes and data saturation was assessed during the data collection process through frequent revision of the probes.

HCPs engaged in two steps. Firstly, they reviewed 2 transcripts and summaries of interactions between CMs and *Your Path*. Secondly, they participated in semi-structured interviews, each lasting approximately 30 minutes, where they assessed the clinical appropriateness of *Your Path* by examining whether it provided clear and accurate guidance on next steps after an HIV self-test, offered emotional support, and facilitated conversations about post-test counselling. They also provided feedback on the completeness and relevance of the AI-generated summaries of the same transcripts.

### Data Analysis

Statistical analyses were conducted using Stata 18 [20]. Descriptive statistics, including percentages, means, medians and standard deviations, were used to summarise socio-demographic variables and survey responses. CMs’ perception of the usability of *Your Path* was assessed using the SUS, a validated 10-item questionnaire in which each item is rated on a 5-point Likert scale [21]. A mean SUS score of 68 or higher is considered above average, while a score of 80 or above indicates excellent usability. However, interpretation of SUS scores can vary based on the system’s complexity, target audience, and context of use.

Qualitative data was analysed using NVivo version 14. Audio recordings were transcribed using a professional transcription service. Thematic analysis using a deductive approach was used to analyse the data. Codebooks were developed and refined through an iterative approach by three researchers (NM, CG & OM). During discussions, inter-rater reliability across themes were assessed where similar themes were retained while others were dropped when no consensus was reached. The study results were aligned with the Theoretical Framework of Acceptability (TFA) [22].

## RESULTS

### Sample characteristics

Participants included CMs and HCPS. A total of 154 CMs were invited to participate, of whom 100 were enrolled. Screening failures (n=54) were primarily due to inability to travel to site, work schedule conflicts and a lack of interest in participating. Similarly, 37 HCPs were invited, with 25 enrolled; screen failures (n=12) were largely due to scheduling conflicts or not meeting inclusion criteria. Table 1 presents the sociodemographic characteristics of the enrolled CMs and HCPs. The median age was 28.8 years (IQR 18-61) for CMs and 37.5 years (IQR 24-61) for HCPs. Most CMs were female (59.0%), self-reported HIV-negative (92.0%), and just over half were unemployed (55.0%). Among HCPs, most were female (83.3%), were experienced HIV testing service (HTS) counsellors (60.0%), and 80.8% had been employed for more than three years.

**Table 1:** Community Members and Healthcare Providers sample characteristics (N=125)

| Variables |  | Community Members N=100 (%) | Healthcare Providers N=25 (%) |
| --- | --- | --- | --- |
| **Median Age** |  | 28.8 years (IQR 18-61) | 37.5 years (IQR 24-61) |
| **Gender** | Male | 41 (41.0) | 4 (16.0) |
|  | Female | 59 (59.0) | 21 (84.0) |
| **Self-reported HIV status** | HIV Negative | 92 (92.0) |  |
|  | Did not disclose | 8 (8.0) |  |
| **Employment status** | No | 55 (55.0) |  |
|  | Yes | 45 (45.0) |  |
| **Profession** | Professional Nurse |  | 9 (36.0) |
|  | HTS Counsellor |  | 15 (60.0) |
|  | Health Coach |  | 1 (4.0) |
| **Years of employment in current profession** | Less than 6 months |  | 1 (4.0) |
|  | 1-3 years |  | 6 (24.0) |
|  | 4 years and above |  | 18 (72.0) |

Following the use of *Your Path*, 91.0% of CMs reported increased intention to access HIV services. Over 80.0% of CMs irrespective of their assignment to a positive or negative mock HIV result, recognised the importance of seeking HIV treatment if needed, with 86.7% indicating they were likely to do so, and 78.6% confident in overcoming barriers like cost and time (Online Resource 5). Usability was rated high, with a mean SUS score of 81.6 (SD 17.5) (Table 2). Most CMs found *Your Path* easy to use (83.7%), would use it again (94.9%), and felt confident using it (90.8%). However, 33,6% preferred assistance before using the tool and 15.3% noted that it did not always provide consistent responses to questions where they felt that responses to some queries varied or were unclear.

**Table 2:**
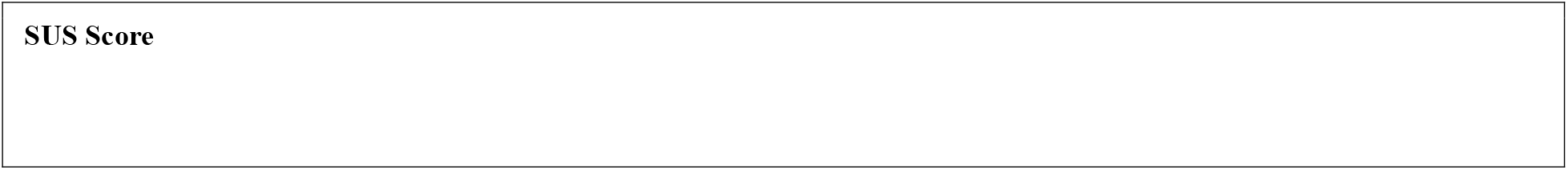
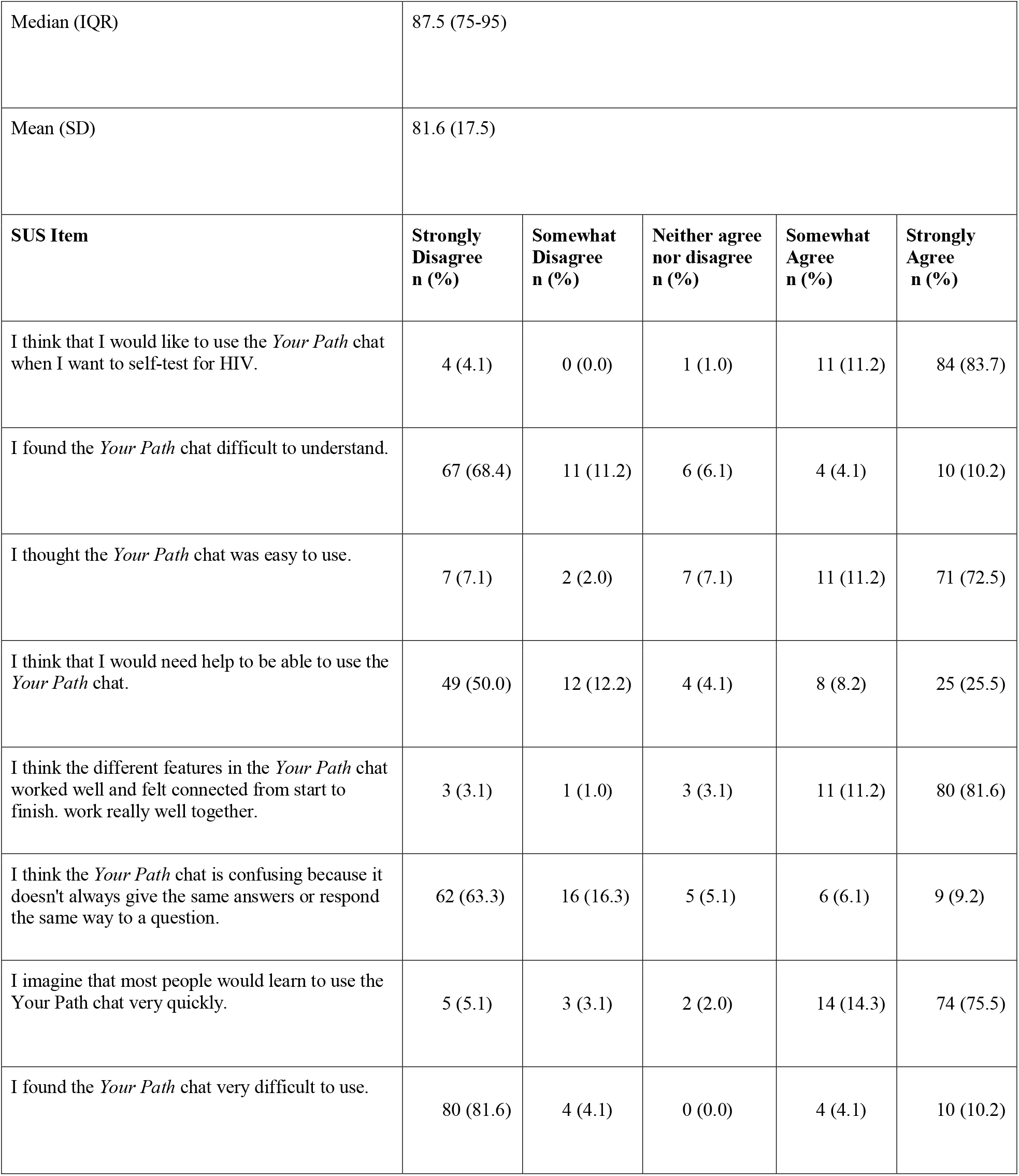
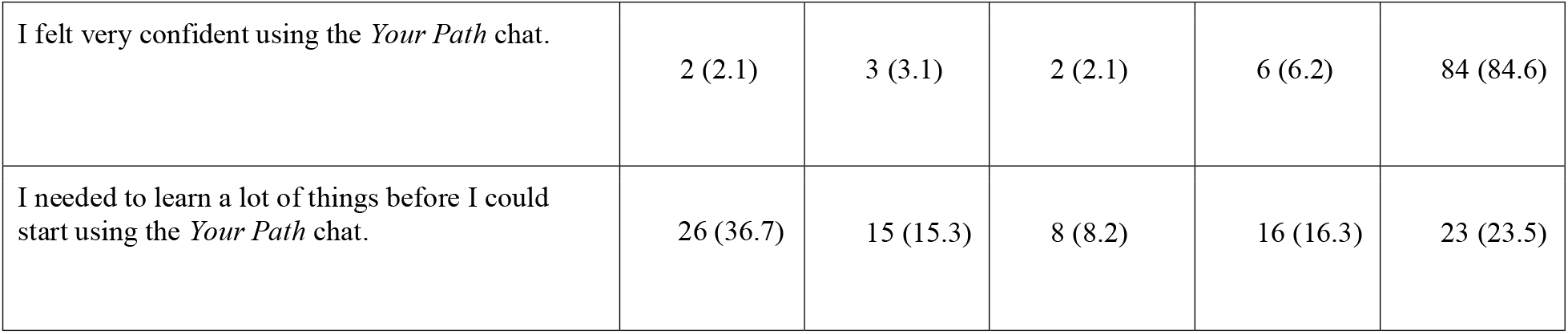
System Usability Scale (SUS) (N=100)

|  |
| --- |
| **SUS Score** |

|  |  |  |  |  |  |
| --- | --- | --- | --- | --- | --- |
| Median (IQR) | 87.5 (75-95) |  |  |  |  |
| Mean (SD) | 81.6 (17.5) |  |  |  |  |
| **SUS Item** | **Strongly Disagree**<br>**n (%)** | **Somewhat Disagree**<br>**n (%)** | **Neither agree**<br>**nor disagree**<br>**n (%)** | **Somewhat Agree**<br>**n (%)** | **Strongly Agree**<br>**n (%)** |
| I think that I would like to use the *Your Path* chat when I want to self-test for HIV. | 4 (4.1) | 0 (0.0) | 1 (1.0) | 11 (11.2) | 84 (83.7) |
| I found the *Your Path* chat difficult to understand. | 67 (68.4) | 11 (11.2) | 6 (6.1) | 4 (4.1) | 10 (10.2) |
| I thought the *Your Path* chat was easy to use. | 7 (7.1) | 2 (2.0) | 7 (7.1) | 11 (11.2) | 71 (72.5) |
| I think that I would need help to be able to use the *Your Path* chat. | 49 (50.0) | 12 (12.2) | 4 (4.1) | 8 (8.2) | 25 (25.5) |
| I think the different features in the *Your Path* chat worked well and felt connected from start to finish. work really well together. | 3 (3.1) | 1 (1.0) | 3 (3.1) | 11 (11.2) | 80 (81.6) |
| I think the *Your Path* chat is confusing because it doesn't always give the same answers or respond the same way to a question. | 62 (63.3) | 16 (16.3) | 5 (5.1) | 6 (6.1) | 9 (9.2) |
| I imagine that most people would learn to use the *Your Path* chat very quickly. | 5 (5.1) | 3 (3.1) | 2 (2.0) | 14 (14.3) | 74 (75.5) |
| I found the *Your Path* chat very difficult to use. | 80 (81.6) | 4 (4.1) | 0 (0.0) | 4 (4.1) | 10 (10.2) |
| I felt very confident using the *Your Path* chat. | 2 (2.1) | 3 (3.1) | 2 (2.1) | 6 (6.2) | 84 (84.6) |
| I needed to learn a lot of things before I could start using the *Your Path* chat. | 26 (36.7) | 15 (15.3) | 8 (8.2) | 16 (16.3) | 23 (23.5) |

The deductive themes developed in accordance with the Theoretical Framework of Acceptability (TFA), and derived from the in-depth interviews are presented below and summarised in Table 3.

**Table 3:** Deductive themes aligned with Theoretical Framework of Acceptability.

| **Affective Attitude** | **Burden** | **Ethicality** | **Intervention coherence** | **Perceived effectiveness** |
| --- | --- | --- | --- | --- |
| Feelings of Community members when engaging with *Your Path* and emotions expressed by healthcare providers when reviewing the transcripts | Community members and healthcare providers concerns about *Your Path* | Community members comparison of *Your Path* with healthcare provider interactions | Community members' understanding and ability to use *Your Path* | Community members and healthcare providers perceived benefits of *Your Path* |

*Source: Adapted from Sekhon et al, 2017*

### Feelings expressed when engaging with *Your Path* and reviewing transcripts [Affective Attitude]

CMs described the interactions with *Your Path* as warm, welcoming, and offering personalised support that was comforting and reassuring.

*“It felt like I was talking to somebody. So, for me I felt like the app, you talking to another human being … like it was so warm, it was full of emotions*.*”* (Female, aged 26-35, negative mock results)

*“It made me feel better. You know that as a man it is not easy to talk to people. Your Path made things simpler*.*”* (Male, aged 26-35, positive mock results)

However, when HCPs were reviewing the transcripts and summaries, they felt that *Your Path* did not provide adequate pre-counselling and the interaction seemed robotic, lacking the warmth and responsiveness of human interaction.

*“When you do counselling with a patient, you become more empathetic with the patient, so when you’re using the chatbot you cannot reach that level with the patient. So, when it’s face to face and you’re there with the patient*,

*you can see their emotions through their eyes and their body language. When using the chatbot you can’t. You just answer whatever that they are texting*.*”* (Female, aged 36-45)

*“I feel because it’s, it’s AI man, it’s impersonal. It doesn’t give an opportunity to … the client to express their feelings…”* (Male, aged 36-45)

### Concerns expressed about *Your Path* [Burden]

CMs and HCPs raised concerns about privacy, language limitations, and overall accessibility of *Your Path*.

*“*…*I don’t believe that there will be privacy or something…I had the concern, that one can see it …”* (Male, aged 36-46, negative mock results)

*“I think they should add more pictures and be able to use the vernacular, …because some people do not know English that well*.*”* (Female, aged 36-45)

*“Only a few things people would complain about is data…*..*and maybe access to Wi Fi*.*”* (Male, aged 36-45)

### Comparison of a digital platform versus in-person interactions for counselling [Ethicality]

CMs perceived interactions with *Your Path* as friendly, and non-judgmental when compared to engaging with HCPs during counselling sessions.

*“*…*I won’t get any judgmental features, I can say whatever … and it won’t give me any judgment at the end of the day… People are judgemental*…*It is much better with Your Path*.*”* (Male, aged 18-25, negative mock results)

*“When being answered there, like I was not being judged … if they had said I should go to the clinic, those questions that I was asking through the Your Path, I don’t see, I would have asked them, a sister like while looking at her cause hey! They judge you*.*”* (Female, aged 36-45, positive mock results)

### Community member’s ability to understand and to use *Your Path* [Intervention coherence]

CMs described *Your Path* as user-friendly, simple and convenient to use, enabling straightforward access to information and support.

*“So, much simple, it is simple this thing…it responds and quick*.*”* (Female, aged 46+, positive mock results)

*“*…*It’s much easier to use … Al, so it’s so much easier for you to understand what it refers you to and what it says to you, I think, with your direct question*.*”* (Male, aged 18-25, positive mock results)

### Perceived benefits of *Your Pat*h for HIV services [Perceived effectiveness]

CMs and HCPs felt that *Your Path* provided useful and relevant step-by-step guidance for HIV self-testing and appropriate information on HIV prevention and treatment supporting linkage to appropriate HIV services. HCPs also viewed it as a helpful tool for individuals hesitant to visit clinics.

*“I gained a lot of knowledge…I didn’t know about PrEP. That PrEP are pills that prevent you from getting HIV*, …*I didn’t know that if you take your medication well, while positive, you are able to not infect other people…”* (Female, aged 18-26, positive mock results)

*“What I liked is that the AI… was talking to the participant that he must make sure he goes to the nearest clinic to confirm the result that he or she got …”* (Female, aged 36-45)

*“There are people who will need help, but they’re afraid because of stigma. So, I think this AI is a good thing, because a person can go to her room or his room alone and do this, and then they know their status … if they listen to the AI to go and look for assistance at the nearest clinic*.*”* (Female, aged 36-45)

## DISCUSSION

In this exploratory study conducted in a controlled setting, we found that *Your Path*, an AI-powered tool designed to facilitate linkage to HIV prevention and treatment services following HIV self-testing (HIVST), is usable, acceptable, and clinically appropriate. To our knowledge, this study is among the first conducted in a resource-limited setting to evaluate a digital AI-powered technology specifically focused on strengthening post-HIVST linkage to care, highlighting the potential of AI-powered interventions to address gaps in HIV service delivery. These findings underscore the promise of user-centered digital tools to support timely connection to care, particularly in contexts where access to healthcare resources is constrained.

Most studies that evaluated the use of digital tools for public health services have been conducted in high-income countries with well-established digital infrastructure [23, 24, 25, 26]. However, recent research in Africa has begun to explore the role of mobile health (mHealth) interventions, such as SMS reminders, and chat-based platforms in improving HIV testing uptake, medication adherence, and clinic appointment reminders [27, 28, 29, 30, 31,32, 33]. Despite progress in research evaluating the integration of digital tools into public health, few studies have specifically examined how AI-powered conversational agents can support and strengthen linkage to HIV prevention and treatment services in South Africa. Our study contributes to closing this gap by evaluating the usability, acceptability, and clinical appropriateness of the AI-powered *Your Path* tool, designed to facilitate linkage to HIV prevention and treatment services following HIV self-testing in South Africa. Most studies exploring the use of digital tools in public health focused primarily on the perspective of clients [25, 26, 28,29,47,48] while our study included both the perspectives of clients and HCPs to capture a comprehensive understanding.

Within the HIV programme, linkage to care following an HIVST is poor and this is attributed to behavioural barriers such as stigma and lack of guidance on the next steps [34, 35]. AI tools present an opportunity to address these barriers, particularly through application in digital conversational agents. Studies have shown that AI-based conversational agents are acceptable within an HIV context, and can engage users on the next steps post HIVST in a manner that is private, confidential and non-judgemental [28]. For instance, a study conducted by Garett R, Young shows that these features make conversational agents especially valuable for high-risk populations, offering stigma free entry points into HIV services [37]. Our study findings align with this, where CMs expressed a preference for engaging with the AI companion over a real counsellor due to fear of stigma, judgement and perceived lack of confidentiality with the human interaction. These findings are also consistent with results from another study [38], where users often preferred digital healthcare agents over human providers when discussing sensitive issues.

Our findings indicate that *Your Path* is acceptable, which aligns with studies conducted in Malaysia [35] and South Africa [39, 28, 40] as well as a broader scoping review [41], which show that digital tools integrated with AI are particularly well-received in the context of HIV care. Acceptability of digital interventions is influenced by multiple factors, including usability, confidentiality, and emotional support. In our study, there was high willingness to use *Your Path*, reflecting openness to incorporating digital technologies into HIV self-testing and post-test support.

Qualitative interviews highlighted emotional safety and confidentiality as key contributors to user satisfaction. Many people living with HIV still face significant psychological challenges as they are often discriminated against, socially isolated, and stigmatised for their condition [44, 45, 46]. Our findings, along with those of other studies [39, 45], suggest that a non-judgmental, anonymous conversational agent like *Your Path* can provide a reassuring and safe space for users to discuss sensitive topics, such as HIV self-testing results without fear of being criticised. This may encourage users to seek information freely and might enhance their willingness to use *Your Path*.

Our findings also highlight strong usability of *Your Path*, with a mean usability score (81.6/68) exceeding the pre-set threshold for success. Participants reported high ease of use and described the tool as accessible and simple to navigate, consistent with other studies showing the value of digital interventions when designed for familiarity and convenience [45, 46]. The predominance of young participants in our sample may have contributed to these positive perceptions, given the widespread use of WhatsApp and uptake of mobile technology among youth, consistent with previous studies [28, 47, 48]. Together, these findings suggest that *Your Path* is well-positioned to engage users by combining high acceptability with strong usability, both of which are critical for sustaining adoption of digital health tools in HIV care.

### Strengths and limitations

Several concerns and limitations, including data privacy, security of personal information, and data access were highlighted despite positive perceptions of *Your Path*, a finding consistent with other studies on digital health interventions in sub-Saharan Africa [49, 50, 51]. To align with the South African POPI Act (2013) [52] and strengthen user trust, future versions of *Your Path* could implement explicit user consent processes, minimize collection of personal identifiers, and provide users with the option to delete their data. The reliance of the current version of *Your Path* on mobile data and the exclusive use of English are limitations that could hinder accessibility, particularly for users in low-connectivity areas or those not proficient in English. Addressing these challenges may require offering data-free access, a strategy already used in South African mHealth programs such as Mom Connect, B-Wise, and the Moya app, which expanded reach by eliminating data costs for end-users [53, 54, 55]. To address language limitations, future versions of *Your Path* could include other local and regional languages to increase reach and access.

This study has contributed important information to the acceptability, usability and clinical appropriateness of AI-powered tools for HIV care in South Africa, but it has limitations. Recruitment was limited to a single urban site, where participants were more likely to have access to and be comfortable with digital technologies. As a result, our findings may not be fully generalisable to rural populations where access to and familiarity with such technologies may differ. We conducted our study in a controlled simulated environment, which means, the actual linkage to care was not measured, and reported intentions to link to care may have been influenced by social desirability bias. However, the simulated environment was necessary to allow for safe, ethical testing of feasibility before moving to real-world implementation. Furthermore, our study sample was skewed towards younger adults who may be more comfortable with digital tools, potentially underrepresenting older or less tech-savvy individuals. The limitations in our study underscore the importance of future field-based evaluation with a more diverse population. Such studies will be essential to assess the acceptability, usability and clinical appropriateness of *Your Path* in a real-life context where logistical, behavioural, and system-level factors play a larger role. Strengths of this study include its mixed-methods design which allowed for a comprehensive, user-centered evaluation of acceptability, usability, and clinical appropriateness. By incorporating direct input from both community members and healthcare providers, the study ensured that *Your Path* was evaluated in alignment with the real-world needs and preferences of its end users.

### Conclusions

In conclusion, this study found *Your Path* to be usable, acceptable and clinically appropriate as a digital health tool to support linkage to services after an HIVST. Participants valued its ability to provide convenient access to accurate information, step-by-step guidance, and personalised support for connecting with HIV services. Insights from this study will inform future builds and adaptations of *Your Path* to enhance user experience, improve integration within health systems, and ensure responsiveness to emerging needs in HIV prevention and treatment. Future studies should evaluate *Your Path* in clinical settings to determine its real-world effectiveness, and conduct cost-effectiveness analyses to assess its value, scalability and sustainability of AI-powered digital health tools for supporting HIV care and broader health service delivery.

## Declarations

### Ethics approval

The Wits Human Research Ethics Committee (Medical) (Reference number 211122) approved the study, and was conducted in accordance with the ethical standards laid down in the 1964 Declaration of Helsinki and its later amendments or comparable ethical standards.

### Consent to participate

Written informed consent was obtained from all participants prior to their participation in the study.

### Data Availability

The manuscript has no associated data in a public repository. The datasets generated and/or analysed during the current study are not publicly available due to confidentiality agreements with participants but are available from the corresponding author on reasonable request.

### AI Disclosure

The authors used OpenAI’s ChatGPT to assist with language refinement and grammar checking to improve the clarity and readability of the manuscript. All intellectual content, data analysis, and interpretation were conducted solely by the authors, who take full responsibility for the final version of the manuscript.

## Notes

### Competing Interest Statement

The authors have declared no competing interest.

### Funding Statement

The research reported in this publication was supported by the South African Medical Research Council (SAMRC) with funds received from the National Department of Health and the Gates Foundation. LL was supported by the National Institute of Mental Health of the National Institutes of Health under grant number K01MH119923. The content and findings reported/illustrated are the sole deduction, view, and responsibility of the researchers and do not reflect the official position and sentiments of the SAMRC, Gates Foundation or the National Institutes of Health.

