## Supplementary Materials for "Evaluating the acceptability, usability and clinical appropriateness of *Your Path*, an AI-powered tool facilitating relevant access to HIV services post-HIV self-testing in South Africa"

| **Online Resource 1: Table 1A. Consolidated criteria for reporting qualitative research (COREQ) checklist** | | | |
| --- | --- | --- | --- |
| **Topic** | **Item No** | **Guide Questions/Description** | **Reported on**  **Page No.** |
| **Domain 1: Research team**  **and reflexivity** | | | |
| Personal characteristics | | | |
| Interviewer/facilitator | 1 | Which author/s conducted the interview or focus group | 5 |
| Credentials | 2 | What were the researcher’s credentials? E.g. PhD, MD | 5 |
| Occupation | 3 | What was their occupation at the time of the study? | 5 |
| Gender | 4 | Was the researcher male or female? | 5 |
| Experience and training | 5 | What experience or training did the researcher have | 5 |
| Relationship with  participants | | | |
| Relationship established | 6 | Was a relationship established prior to study commencement? | N/A |
| Participant knowledge of the interviewer | 7 | What did the participants know about the researcher? e.g. personal  goals, reasons for doing the research | N/A |
| Interviewer characteristics | 8 | What characteristics were reported about the interviewer/facilitator?  e.g. Bias, assumptions, reasons and interests in the research topic | N./A |
| **Domain 2: Study design** | | | |
| Theoretical framework | | | |
| Methodological orientation and Theory | 9 | What methodological orientation was stated to underpin the study? e.g.  grounded theory, discourse analysis, ethnography, phenomenology,  content analysis | 4 |
| Participant selection | | | |
| Sampling | 10 | How were participants selected? e.g. purposive, convenience,  consecutive, snowball | 4 |
| Method of approach | 11 | How were participants approached? e.g. face-to-face, telephone, mail,  email | 4 |
| Sample size | 12 | How many participants were in the study? | 4 |
| Non-participation | 13 | How many people refused to participate or dropped out? Reasons? | 6 |
| Setting | | | |
| Setting of data collection | 14 | Where was the data collected? e.g. home, clinic, workplace | 4 |
| Presence of nonparticipants | 16 | Was anyone else present besides the participants and researchers? | 4 |
| Description of sample | 17 | What are the important characteristics of the sample? e.g. demographic data, date | 6 |
| Data collection | | | |
| Interview guide | 18 | Were questions, prompts, guides provided by the authors? Was it pilot tested? | 5 |
| Repeat interviews | 1 | Were repeat interviews carried out? If yes, how many? | N/A |
| Audio/visual recording | 19 | Did the research use audio or visual recording to collect the data? | 5 |
| Field notes | 20 | Were field notes made during and/or after the interview or focus group? | N/A |
| Duration | 21 | What was the duration of the interviews or focus group? | 5 |
| Data saturation | 22 | Was data saturation discussed? | 5 |
| Transcripts returned | 22 | Were transcripts returned to participants for comment and/or correction | N/A |
| **Domain 3: Data analysis and findings** | | | |
| **Number of data coders** | 24 | How many data coders coded the data? | 5 |
| Description of the coding  tree | 25 | Did authors provide a description of the coding tree? | N/A |
| Derivation of themes | 26 | Were themes identified in advance or derived from the data? | 6 |
| Software | 27 | What software, if applicable, was used to manage the data? | 6 |
| Participant checking | 28 | Did participants provide feedback on the findings? | N/A |
| Reporting | | | |
| Quotations presented | 29 | Were participant quotations presented to illustrate the themes/findings? | 6 - 8 |
| Data and findings consistent | 30 | Was there consistency between the data presented and the findings? | 6 - 8 |
| Clarity of major themes | 31 | Were major themes clearly presented in the findings? | 6 - 8 |
| Clarity of minor themes | 32 | Is there a description of diverse cases or discussion of minor themes? | N/A |

Source: Tong A, Sainsbury P, Craig J. Consolidated criteria for reporting qualitative research (COREQ): a 32-item checklist for interviews and focus groups. International Journal for Quality in Health Care. 2007. Volume 19, Number 6: pp. 349 – 357

| **Online Resource 2: Table 1B. Pre-test survey to measure intention to seek HIV services among community members** | | | | | | |
| --- | --- | --- | --- | --- | --- | --- |
| **Scales for participants who receive an image of a HIV negative mock test result** | | | | | | |
| Please read the following statement and tick only one box for the statement as to how much you agree or disagree with the statement based on your mock HIV self-test results that you have received. | | | | | | |
|  | **Strongly disagree** | **Somewhat Disagree** | **Neither agree nor disagree** | **Somewhat agree** | **Strongly agree** | |
| Engaging in HIV prevention services is beneficial for my health. |  |  |  |  |  | |
| Seeking HIV treatment services is important if I test positive. |  |  |  |  |  | |
| Please read the statement below and tick only one box for the statement as to how confident you are about your ability to access treatment services based on your mock HIV self-test results that you have received. | | | | | | |
|  | **Not confident at all** | **Not confident** | **Unsure** | **Confident** | **Very confident** | |
| I am confident that I can access HIV prevention services if I need to. |  |  |  |  |  | |
| I am confident that I can find the resources (e.g. money and time) to seek HIV treatment services if I test positive. |  |  |  |  |  | |
| Please read the question below and tick only one box for the question as to how likely you think you would be to seek HIV prevention and future HIV treatment services based on your mock HIV self-test result that you have received. | | | | | | |
|  | **Extremely likely** | **Very likely** | **Somewhat likely** | **Somewhat unlikely** | **Very unlikely** | **Extremely unlikely** |
| How likely are you to engage in HIV prevention services? |  |  |  |  |  |  |
| How likely are you to seek future HIV treatment services if you test HIV positive? |  |  |  |  |  |  |
| **Scales for participants who receive an image of an HIV positive mock test result** | | | | | | |
| Please read the following statement and tick only one box for the statement as to how much you agree or disagree with the statement based on your mock test result that you have received | | | | | | |
|  | **Strongly disagree** | **Somewhat Disagree** | **Neither agree nor disagree** | **Somewhat agree** | **Strongly agree** | |
| Seeking HIV treatment services is important. |  |  |  |  |  | |
| Please read the statement below and tick only one box for the statement as to how confident you are about your ability to access treatment services based on the mock test result that you have received. | | | | | | |
|  | **Not confident at all** | **Not confident** | **Unsure** | **Confident** | **Very confident** | |
| I am confident that I can find the resources (e.g. money and time) to seek HIV treatment services. |  |  |  |  |  | |
| Please read the question below and tick only one box for the question as to how likely you think you would be to seek HIV treatment services based on your mock HIV test results. | | | | | | |
|  | **Extremely likely** | **Very likely** | **Somewhat likely** | **Somewhat unlikely** | **Very unlikely** | **Extremely unlikely** |
| How likely are you to seek HIV treatment services? |  |  |  |  |  |  |

| **Online Resource 3: Table 1C. Post-test survey to measure intention to seek HIV services and Your Path usability** | | | | | | |
| --- | --- | --- | --- | --- | --- | --- |
| **Scales for participants who receive an image of an HIV negative mock test result** | | | | | | |
| Please read the following statements and tick only one box for the statement as to how much you agree or disagree with the statements based on your mock HIV self-test results that you have received. | | | | | | |
|  | **Strongly disagree** | **Somewhat Disagree** | **Neither agree nor disagree** | **Somewhat agree** | **Strongly agree** | |
| Engaging in HIV prevention services is beneficial for my health. |  |  |  |  |  | |
| Seeking HIV treatment services is important if I test positive. |  |  |  |  |  | |
| Please read the statements below and tick only one box for the statements as to how confident you are about your ability to access treatment services based on your mock HIV self-test results that you have received. | | | | | | |
|  | **Not confident at all** | **Not confident** | **Unsure** | **Confident** | **Very confident** | |
| I am confident that I can access HIV prevention services if I need to. |  |  |  |  |  | |
| I am confident that I can find the resources (e.g. money and time) to seek HIV treatment services if I test positive. |  |  |  |  |  | |
| Please read the questions below and tick only one box for the questions as to how likely you think you would be to seek HIV prevention and future HIV treatment services based on your mock HIV self-test result that you have received. | | | | | | |
|  | **Extremely likely** | **Very likely** | **Somewhat likely** | **Somewhat unlikely** | **Very unlikely** | **Extremely unlikely** |
| How likely are you to engage in HIV prevention services? |  |  |  |  |  |  |
| How likely are you to seek future HIV treatment services if you test HIV positive? |  |  |  |  |  |  |

| Please read each statement below and tick only one box for each statement as to whether you agree with the statement or not about the usability of the “Your Path” chat. | | | | | |
| --- | --- | --- | --- | --- | --- |
|  | **Strongly Disagree** | **Somewhat Disagree** | **Neither agree nor disagree** | **Somewhat Agree** | **Strongly Agree** |
| I think that I would like to use the “Your Path” chat when I want to self-test for HIV. |  |  |  |  |  |
| I found the “Your Path” chat difficult to understand. |  |  |  |  |  |
| I thought the “Your Path” chat was easy to use. |  |  |  |  |  |
| I think that I would need help to be able to use the “Your Path” chat. |  |  |  |  |  |
| I think the different features in the "Your Path" chat worked well and felt connected from start to finish. work really well together. |  |  |  |  |  |
| I think the “Your Path” chat is confusing because it doesn't always give the same answers or respond the same way to a question. |  |  |  |  |  |
| I imagine that most people would learn to use the “Your Path” chat very quickly. |  |  |  |  |  |
| I found the “Your Path” chat very difficult to use. |  |  |  |  |  |
| I felt very confident using the “Your Path” chat. |  |  |  |  |  |
| I needed to learn a lot of things before I could start using the “Your Path” chat. |  |  |  |  |  |
| **Additional questions** | 1. Did interaction with the Your Path chat influence your decision to access HIV prevention or future HIV treatment services? | | | | |
|  | 2. Please provide an explanation on how your interaction with the Your Path chat influence your intentions to access HIV care? | | | | |
| **Scales for participants who receive an image of an HIV positive mock test result** | | | | | |
| Please read the following statement and tick only one box for the statement as to how much you agree or disagree with the statement based on your mock test result that you have received**.** | | | | | |
|  | **Strongly disagree** | **Somewhat Disagree** | **Neither agree nor disagree** | **Somewhat agree** | **Strongly agree** |
| Seeking HIV treatment services is important |  |  |  |  |  |
| Please read the statement below and tick only one box for the statement as to how confident you are about your ability to access treatment services based on the mock test result that you have received. | | | | | |
|  | **Not confident at all** | **Not confident** | **Unsure** | **Confident** | **Very confident** |
| I am confident that I can find the resources (e.g. money and time) to seek HIV treatment services. |  |  |  |  |  |
| Please read the question below and tick only one box for the question as to how likely you think you would be to seek HIV treatment services based on your mock HIV test results that you have received. | | | | | |
|  | **Extremely likely** | **Very likely** | **Somewhat likely** | **Somewhat unlikely** | **Very unlikely** |
| How likely are you to seek HIV treatment services? |  |  |  |  |  |
| Please read each statement below and tick only one box for each statement as to whether you agree with the statement or not about the usability of the “Your Path” chat. | | | | | |
|  | **Strongly Disagree** | **Somewhat Disagree** | **Neither agree nor disagree** | **Somewhat Agree** | **Strongly Agree** |
| I think that I would like to use the “Your Path” chat when I want to self-test for HIV. |  |  |  |  |  |
| I found the “Your Path” chat difficult to understand. |  |  |  |  |  |
| I thought the “Your Path” chat was easy to use. |  |  |  |  |  |
| I think that I would need help to be able to use the “Your Path” chat. |  |  |  |  |  |
| I think the different features in the "Your Path" chat worked well and felt connected from start to finish. work really well together. |  |  |  |  |  |
| I think the “Your Path” chat is confusing because it doesn't always give the same answers or respond the same way to a question. |  |  |  |  |  |
| I imagine that most people would learn to use the “Your Path” chat very quickly. |  |  |  |  |  |
| I found the “Your Path” chat very difficult to use. |  |  |  |  |  |
| I felt very confident using the “Your Path” chat. |  |  |  |  |  |
| I needed to learn a lot of things before I could start using the “Your Path” chat. |  |  |  |  |  |
| **Additional questions** | 1. Did interaction with the Your Path chat influence your decision to access HIV prevention or future HIV treatment services? | | | | |
|  | 2. Please provide an explanation on how your interaction with the Your Path chat influence your intentions to access HIV care? | | | | |

### **Online Resource 4: Interview guide/prompts for Community Members**

1. **How would you describe “Your Path” to someone who has never used it before?**

*Probe:*

- *When you think about “Your Path”, what first comes to mind?*
- *What do you remember about using “Your Path”?*
- *Did anything stand out to you?*
- *How did using “Your Path “make you feel? Why do you think it made you feel this way?*
- *How would you describe the "personality" of “Your Path”?*

2. **What was the most interesting aspect of “Your Path” for you?**

*Probe:*

- *What caught your attention?*
- *What was different about this interaction and similar discussions with a nurse/clinician/doctor?*

3. **Can you tell me about your experience using "Your Path" compared to talking to someone about the same issues? What did you find most appealing?**

*Probe:*

- *Was there a sense of security or privacy that you felt more with “Your Path” than with a person?*
- *Did you find it to be less judgmental compared to talking to a human?*
- *Did you feel a sense of connection or care from “Your Path”?*

4. **Did you encounter any challenges or areas for improvement while using "Your Path"?**

*Probe:*

- *Were there any aspects of “Your Path” that you found confusing?*
- *Did anything about it make you feel frustrated or upset?*

5. **Where would you ideally hear about “Your Path” and its counselling features?**

*Probe:*

- *Radio*
- *Billboards*
- *Local health clinics*
- *Health care worker*
- *Friend or family*
- *Somewhere else*

6. **What do you think happens with the information that you enter into “Your Path”?**

*Probe:*

- *What are your concerns about data privacy?*
- *Do you think anything should be shared with a clinician after you finish chatting with “Your Path”? e.g., a summary, the whole conversation?*
- *How would you feel if a nurse saw a summary of your conversation?*
- *How would you feel if the whole conversation were provided to a nurse to follow-up with you about accessing health services after an HIV self-test?*

7. **Can you imagine other issues where using a chat like ‘Your Path’ for a conversation would be useful outside of HIV self-testing?**

*Probe:*

- *Would you like to use an experience like this to access other health information?*
- *Would you like to use an experience like this to access information about your health or symptoms when you’re feeling ill?*

8. **If you had a choice of discussing a difficult topic like your HIV self-test result with either a healthcare provider in person or with “Your Path,” which would you choose and why?**
9. **What would you change about “Your Path”?**

*Probe:*

- *Was anything confusing or unclear?*
- *Too much information?*
- *Was this information you already knew?*
- *Was it boring?*
- *Was it too invasive or felt too personal?*
- *Did you have any questions for “Your Path”? If so, did you get to ask them? Did you get the answers that you needed?*

10. **Did “Your Path” change your thinking at all about the benefits of accessing health services after an HIV self-test?**

*Probe:*

- *Do you think it is important to know the next steps for engagement with health services after an HIV self-test?*

11. **Do you think other people would benefit from using “Your Path”?**

*Probe*:

- *Who would most benefit from the use of something like “Your Path?”*

12. **Any final thoughts on using “Your Path”?**

| **Online Resource 5: Table 1D. Intention to seek HIV services among CMs (N=100)** | | | |
| --- | --- | --- | --- |
|  | | **Pre-Test Survey**  **N (%)** | **Post-Test Survey**  **N (%)** |
| **Seeking HIV treatment services is important.** | Strongly disagree | 12 (12.0) | 10 (10.2) |
|  | Somewhat Disagree | 3 (15.0) | 0 (0.0) |
|  | Neither agree nor disagree | 3 (3.0) | 3 (3.1) |
|  | Somewhat agree | 4 (4.0) | 1 (1.0) |
|  | Strongly agree | 78 (78.0) | 84 (85.7) |
| **I am confident that I can find the resources (e.g. money and time) to seek HIV treatment services.** | Not confident at all | 7 (7.0) | 5 (5.1) |
|  | Not confident | 7 (7,0) | 6 (6.1) |
|  | Unsure | 11 (11.0) | 10 (10.2) |
|  | Confident | 45 (45.0) | 30 (30.6) |
|  | Very confident | 30 (30.0) | 47 (48.0) |
| **How likely are you to seek HIV treatment services?** | Extremely unlikely | 6 (6.0) | 5 (5.1) |
|  | Very unlikely | 7 (7.0) | 4 (4.1) |
|  | Somewhat unlikely | 6 (6.0) | 4 (4.1) |
|  | Very likely | 30 (30.0) | 27 (27.6) |
|  | Extremely likely | 51 (51.0) | 58 (59.2) |
